# Pertussis upsurge, age shift and vaccine escape post-COVID-19 caused by *ptxP3* macrolide-resistant *Bordetella pertussis* MT28 clone in China: a genomic epidemiology study

**DOI:** 10.1101/2024.04.16.24305932

**Authors:** Pan Fu, Gangfen Yan, Yijia Li, Li Xie, Yuehua Ke, Shuxiang Qiu, Shuang Wu, Xiaolu Shi, Jie Qin, Jinlan Zhou, Guoping Lu, Chao Yang, Chuanqing Wang

## Abstract

**Background:** The upsurge of pertussis post-COVID-19 and expansion of macrolide-resistant *Bordetella pertussis* (MRBP) pose significant public health challenges worldwide. China has experienced notable pertussis upsurge post-COVID-19, alongside an age shift to older children, vaccine escape and a notable rise in MRBP prevalence. We describe the genomic epidemiological investigation of these events.

**Method:** We did a retrospective, population-based study using culture-positive *B. pertussis* from Children’s Hospital of Fudan University (CHFU), the exclusive referral hospital for childhood notifiable infectious diseases, in Shanghai, China between June 2016 and March 2024. We analysed strain and pertussis epidemiology dynamics by integrating whole-genome sequencing of 723 strains with antimicrobial susceptibility, transcriptomic proflie, and clinical data. We compared the genome sequences of Shanghai strains with 6450 Chinese and global strains.

**Findings:** Coincident with national situtation, pertussis cases upsurged post-COVID-19 in Shanghai. At CHFU, the number of confirmed cases (n=349) in the first three months of 2024 exceeded the total case of previously years (n≤177). Post-COVID-19, patients shifted from predominantly infants (90%, 397/442) to widespread infection among older children (infant: 16%, 132/844), with vaccinated individuals surging from 31% (107/340) to 88% (664/756); MRBP prevalence increased from 60% (267/447) to 98% (830/845). The emergence and expansion of a *ptxP3*-linage, macrolide-resistant novel clone with MLVA type 28, MR-MT28, uniquely capable of causing substantial infections among older children and vaccinated individuals, temporally strongly associated with the pertussis upsurge and epidemiological transition. MR-MT28 exhibited increased expression of antigen genes including pertussis toxin genes, along with high incidence of abnormal C-reactive protein, but associated with siginicantly milder clinical symtoms (e.g. wheezing, facial blushing, *p*<0·01), higher proportion of normal chest computed tomography (*p*<0·05) and lower hospitalization rate (*p*<0·01). Phylogenomic clustering analysis revealed a higher proportion of MR-MT28 strains grouping into clusters representing putative transmission. We reconstructed the evolutionary history of MR-MT28, and showed that it most likely originated in China around 2016 (95% highest probability density: 2013-2017) after acquring several mutations, including a novel antigen allele *prn150* and 23S rRNA A2047G mutation. Approximately one quarter (26%, 50/195) of MR-MT28 has evolved into predicted PRN-deficient strains. MR-MT28 has been identified in four regions (Anhui, Shanghai, Beijing and Guangdong) of China and continuously detected in Shanghai and Beijing, suggesting domestic spread and colonization.

**Interpretation:** We identified a *ptxP3*-linage, macrolide-resistant novel clone, MR-MT28, and provide evidence that pathogen evolution is more likely the primary factor driving pertussis upsurge, age shift and vaccine escape. MR-MT28 potentially poses a high global spread risk and warrants global surveillance. Macrolides may no longer be suitable as first-line drugs for pertussis treatment in China.

**Funding:** National Key Research and Development Program of China (2021YFC2701800 and 2022YFC2304700), National Natural Science Foundation of China (82202567 and 32270003), Youth Innovation Promotion Association, Chinese Academy of Sciences (2022278), Shanghai Rising-Star Program (23QA1410500), and Shanghai municipal three-year action plan for strengthening the construction of the public health system (2023-2025) GWVI-2.1.2.

**Research in context:** *Evidence before this study:* In the first two months of 2024, an unexpected upsurge in pertussis was seen in both China and Europe. Furthermore, the pertussis upsurge in China exhibited atypical patterns, including an age shift to older children, vaccine escape and a notable increase in macrolide-resistant *Bordetella pertussis* (MRBP) prevalence. We aimed to test the hypothesis linking pertussis upsurge and epidemiological transition to pathogen evolution. We searched PubMed for molecular epidemiology studies of macrolide-resistant *Bordetella pertussis* using the terms ("*Bordetella pertussis*" OR "pertussis” OR “whooping cough”) AND ("macrolide resistant" OR "erythromycin resistant”) for articles before March 2024 and identified 40 studies. MRBP has been reported in eight counties, including United States, United Kingdom, France, Iran, Cambodia, Vietnam, Japan and China. While MRBP incidence in other countries remained low, it was notably high in China, accounting for 50% and even 90% of strains across various regions. The risk of MRBP spreading out of China was previously considered low, primarily because Chinese strains predominantly belonged to *ptxP1*-lineage, whereas the globally prevalent lineage was *ptxP3*. However, the situation is changing, as *ptxP3*-MRBP strains have been identified in multiple regions of China since 2017. In Shanghai, we identified a sharply increase of *ptxP3*-MRBP prevalence post-COVID-19, coinciding with pertussis age shift to older children and vaccine escape. A similar scenario was independently observed in Beijing. Additionanlly, there is a significant rise in pertussis cases since the beginning of 2024. Currently, there is a lack of study testing the link between pertussis upsurge, epidemiological transition, and the evolution of its causative pathogen.

*Added value of this study:* Our study identified a *ptxP3*-linage, macrolide-resistant novel clone, MR-MT28, which is uniquely capable of causing substantial infections among older children and vaccinated population, suggesting enhanced vaccine escape. The emergence and rapid expansion of MR-MT28 temporally strongly associated with the upsurge of pertussis cases, age shift, vaccine escape and notable rise in MRBP prevalence. MR-MT28 was characterized by increased expression of antigen genes long with high incidence of abnormal C-reactive protein, but associated with siginicantly milder clinical sytmptoms, which may prolong the interval before seeking medical care, thereby amplifying transmission opportunities. Phylogenomic clustering analysis indicated that MR-MT28 may have increased transmissibility. Therefore, MR-MT28 may have competitive advantages due to antimicrobial resistance, enhanced vaccine escape, increased opportunities for transmission and transmissibility. We reconstructed the evolutionary history of MR-MT28 and showed that it most likely originated in China around 2016 after the acquisition of several mutations, and COVID-19 may have promoted its expansion. Approximately one quarter of MR-MT28 strains has evolved into predicted PRN-deficient strains. Our results showed the domestic spread and colonization of MR-MT28.

*Implications of all the available evidence:* Our study provides evidence that pathogen evolution, rather than the widely accepted notion of wanning immunity or ‘immunity debt’, is more likely the primary factor driving pertussis upsurge, age shift and vaccine escape. MR-MT28 potentially poses a high global spread risk, due to its consistent *ptxP3* allele and epidemiology across many counties, together with resistance to first-line drugs and potentially competitive advantages, which warrants global surveillance and research efforts. Macrolides may no longer be suitable as first-line drugs for pertussis treatment in China.

## Introduction

Pertussis (whooping cough) is a highly contagious disease primarily caused by *Bordetella pertussis.* The introduction of the whole-cell vaccines (WCVs) in the 1950s, and the switch to acellular vaccines (ACVs) in the 1980–1990s, significantly reduced pertussis disease burden.^1^ However, the resurge of pertussis has been reported globally during the past two decades.^2–5^ The burden of pertussis is still high, with an estimated 24 million cases and 160,700 deaths in children younger than five years in 2014.^6^ In China, ACV was applied since 2007 and completely replaced WCV in 2012. Despite over 99% vaccine coverage among children, the numbers of reported pertussis cases sharply increased from <3,000 per year in 2006–2013 to 30,027 in 2019.^7^ A reduced incidence of pertussis has been reported in children since the beginning of the COVID-19 pandemic, due to the non-pharmaceutical interventions.^8,9^ However, the recent upsurge in pertussis post-COVID-19 restrictions have been documented in China and Europe.^10–14^ In China, the number of pertussis cases reported in the first two months of 2024 (32,380 cases) was >20 times higher than during the same periods in previous years, approaching the total cases reported for the entire year of 2023 (38,205 cases).^10,11^

Pertussis resurge has been attributed to various factors, including wanning immunity, improved diagnostics, and *B. pertussis* evolution.^2–5^ The importance of *B. pertussis* evolution is suggested by the antigenic divergence between circulating strains and vaccine strains. Sequence divergence has been identified in several genes encoding ACV antigens or their promotor, including filamentous hemagglutinin (Fha), pertactin (Prn), fimbriae (Fim), pertussis toxin (Ptx) and its promoter (*ptxP*).^15^ *ptxP3* strains are currently dominating infections in most of the high-income countries, probably due to their higher virulence caused by more Ptx production.^16^ Moreover, Prn-deficient strains which confer fitness advantages particular to ACV vaccinated populations, have been increasingly reported and became dominant in multiple countries such as Unite States and Australia.^17^ Besides *B. pertussis* evolution toward vaccine escape, the emergence of macrolide-resistant *B. pertussis* (MRBP) mediated by A2047G mutation in the 23s rRNA gene, which confers resistance to first-line drugs such as erythromycin and azithromycin for pertussis treatment, posed further public health issues.^18^

The circulating *B. pertussis* stains and pertussis epidemiology in China differ significantly from those in other countries. *ptxP1* strains have been predominant in China until 2019,^19–21^ rather than globally prevalent *ptxP3* strains. Moreover, the incidence of MRBP in China is notably high, accounting for over 50% and even 90% of strains in various regions,^21–24^ while MRBP remains rare in other countries. Since Chinese MRBP strains were predominantly *ptxP1*, their risk of spreading out of China was previously considered low. Additionally, pertussis patients in China were primarily identified in infants less than one year old,^25^ unlike the other countries where resurgence of pertussis was more observed in older children and adolescents/adults.^4,5^ However, the situation in China is changing, as *ptxP3*-MRBP strains have been identified in multiple regions of China since 2017.^21–24^ In Shanghai, we previously reported that *ptxP3*-MRBP had replaced *ptxP1* strains and dominated infections after 2020, with a substantially increased infections in older, vaccinated children.^22,23^ Similar situation was recently identified in Beijing. During 2021-2022, *ptxP3*-MRBP accounted for 78.8% patients, and the proportion of patients in children over three years old doubled than before.^24^ The *ptxP3*-MRBP potentially poses a high global spread risk because its consistent *ptxP3* allele and epidemiology across many counties.

Coincident with national situtation, pertussis cases upsurged post-COVID-19 in Shanghai. We have carried out continuous surveillance of *B. pertussis* stains and pertussis in Shanghai since 2016, and obtained strains and corresponding clinical data covering the transition period pre- and post-COVID-19. We previously described the genotype dynamics of *ptxP3*-MRBP based on a subset of strains by multi-locus variable-number tandem-repeat analysis (MLVA)^22,23^ Compared with traditional typing methods such as MLVA, whole-genome sequencing (WGS) provides finer resolution and more comprehensive information including virulence and antimicrobial genes. In this study, we took advantages of WGS and sequenced culture-positive *B. pertussis* stains between June 2016 and March 2024. By combining with phenotypic, transcriptomic, and clinical data, we aim to investigate the link between pertussis upsurge, epidemiological transition, and *B. pertussis* evolution. We further compared Shanghai strains with Chinese and global strains to investigate its domestic and global spread risk.

## Methods

### Isolates sampling and participants

Pertussis is a notifiable disease in China. We did a retrospective, population-based study in Shanghai, the most populous city in China, and enrolled pertussis patients from Children’s Hospital of Fudan University (CHFU), the exclusive referral hospital for childhood notifiable infectious diseases. The nasopharyngeal swab samples of patients with suspected pertussis infection are collected and delivered to microbiology laboratory. Pertussis patients were defined as positive culture and/or PCR testing, or showed typical clinical symptoms of pertussis. We studied culture-positive *B. pertussis* between June 2016 and March 2024 (all the culture-positive strains beween June 2016 and August 2023, and randomly selected strains between September 2023 and March 2024, appendix p 2). The laboratory testing results and clinical data were extracted from medical records and all data analysis was anonymous. For comparison, we also included the genomes of 11 randomly selected strains (appendix p 12) collected after 2019 in Beijing (n=8, from Capital Institute of Pediatrics, 2019-2022) and Shenzhen, Guangdong (n=3, from Shenzhen Center for Disease Control and Prevention, 2023). The study protocol was approved by the Ethics Committee of the CHFU (No. 2022-66).

### Whole-genome sequencing and analysis

Genomic DNA of *B. pertussis* strains were extracted using QIAamp DNA mini kit (QIAGEN) and whole-genome sequencing were performed on Illumina NovaSeq platform. Sequencing data were strictly trimmed and analyzed as previously described.^26^ Briefly, species identification based on sequencing data were performed using Kraken 2. Core-genome single-nucleotide-polymorphisms (SNPs) were identified using the Snippy pipeline. Maximum-likelihood phylogenetic trees were constructed using RAxML-NG based on core-genome SNPs. Genome assembly was preformed using shovill pipeline. Assembled sequences were used for vaccine antigen typing by searching against BIGSdb-Pasteur genomic platform for *Bordetella*.^27^ Dated phylogeny and population size dynamics were analyzed using BEAST 1.10 with Skygrid coalescent model. We performed pangenome-level analysis to identified lineage/clone-specific genomic variations, including SNPs, gene presence/absence and unitigs using unitig-caller and pyseer. A total of 6439 publicly available genome from 34 countries and six continents were downloaded from NCBI GenBank or SRA database, with accession numbers listed in the appendix (appendix p 13).

### MLVA and antimicrobial susceptibility testing

Multiple locus variable-number tandem repeat analysis (MLVA) was performed as described by Schouls et al.^28^ The minimum inhibitory concentrations (MICs) of four antimicrobial agents were determined by the E-test. The standardized interpretation criteria are based on our previous report.^22^

### Transcriptomic analysis and quantification of antigen gene expression

The RNA of *B. pertussis* strains was extracted using QIANGEN RNA extraction kit (TIANGEN BIOTECH, Beijing, China). cDNA library construction and sequencing were performed on Illumina NovaSeq platform at Novogene Co. Ltd. (Beijing, China). Differentially expressed genes identification and pathway enrichment analysis were performed using R packages DESeq2 and ClusterProfiler. Transcriptomic and genomic sequencing data have been deposited in the NCBI Sequence Read Archive (SRA) under accession number PRJNA1071282.

To quantify antigen gene expression level, reverse transcription was conducted using YEASEN cDNA Synthesis SuperMix for qPCR kit (Yeasen Biotechnology, Shanghai, China). Amplifications of virulence genes were performed using the Applied Biosystems QuantStudioTM 5 Real-Time qPCR system (ThermoFisher SCIENTIFIC, Waltham, MA, US), with initial denaturation for 5 min at 94 °C, and 35 cycles at 94 °C for 30 s, annealing for 30 s at 52 °C, followed by elongation at 72 °C for 30 s, and a final step at 72 °C for 5 min. The presence of gene specific amplicons was verified visually by 2% agarose gel electrophoresis using Glodview staining. *B. pertussis* ATCC9797 was used as the reference strain and the house-keeping gene *tyrB* was used as the reference gene. The virulence gene expressions were calculated by the comparative CT method (2^-ΔΔCT^).

### Statistical analysis

Datasets were compared with Chi-squared tests for categorical data and Mann-Whitney test for continuous data using GraphPad Prism 9. *P*-values less than 0.05 were considered statistically significant.

## Results

Coincident with national situtation, pertussis cases upsurged post-COVID-19 in Shanghai (appendix p 1). From June 2016 to March 2024, a total of 2659 patients were diagnosed with pertussis in CHFU. Among these, 1479 (56%) patients were confirmed by positive strain cultures, while the others were diagnosed by positive PCR-detection or showed typical pertussis symptoms. We enrolled all the culture-positive *B. pertussis* strains beween June 2016 and August 2023 (n= 714), and 100 randomly selected strains (13% culture-positive strains) between September 2023 and March 2024. A total of 712 (87%) enrolled culture-positive strains were successfully sequenced and identified as *B. pertussis*. The remaining were identified as other *Bordetella* (n=4) or no longer viable (n=98). Of the 712 patients, 678 (95%) were children patients and the remaining were households. Among children patients with available clinical and vaccination data (table 1), 68% (458/672) were infants (aged ≤1y), 46% (310/674) were female, and 52% (284/546) had been vaccinated. A total of 507 (71%) sequenced strains were MRBP, which are resistant to erythromycin, azithromycin and clarithromycin, but all were susceptible to sulfamethoxazole/trimethoprim. All MRBP strains carried the 23S rRNA A2047G mutation.

**Table 1.** Demographic, clinical and laboratory characteristics of children patients.

|  | Total | *ptxP1* | MR-MT28 | MS-MT28 | *ptxP3*-other | *p*-value (MR-MT28 vs others) |
| --- | --- | --- | --- | --- | --- | --- |
| **Age** |  |  |  |  |  |  |
| 0-1y | 458 (68%) | 247 (83%) | 47 (25%) | 21 (100%) | 143 (87%) | All pairs *p*<0.01 |
| >1-3y | 48 (7%) | 20 (7%) | 16 (8%) | / | 12 (7%) |  |
| 4-6y | 77 (11%) | 17 (6%) | 56 (30%) | / | 4 (2%) | All pairs *p*<0.01 |
| 7-10y | 80 (12%) | 13 (4%) | 61 (32%) | / | 6 (4%) | All pairs *p*<0.01 |
| >10y | 9 (1%) | / | 9 (5%) | / | / |  |
| **Gender** |  |  |  |  |  |  |
| Female | 310 (46%) | 136 (45%) | 87 (46%) | 8 (38%) | 79 (48%) |  |
| Male | 365 (54%) | 163 (55%) | 103 (54%) | 13 (62%) | 86 (52%) |  |
| **Vaccination** |  |  |  |  |  |  |
| Yes | 284 (52%) | 89 (39%) | 150 (87%) | 3 (21%) | 42 (33%) | All pairs *p*<0.01 |
| No | 262 (48%) | 142 (61%) | 22 (13%) | 11 (79%) | 87 (67%) | All pairs *p*<0.01 |
| **Clinical characteristics** |  |  |  |  |  |  |
| Spasmodic cough | 593 (87%) | 283 (94%) | 136 (72%) | 20 (95%) | 154 (93%) | All pairs *p*<0.05 |
| Paroxysmal cough | 565 (83%) | 252 (83%) | 159 (84%) | 19 (90%) | 135 (82%) |  |
| Cockcrow-like cough | 123 (18%) | 60 (20%) | 29 (15%) | 3 (14%) | 31 (19%) |  |
| Wheeze | 147 (22%) | 81 (27%) | 14 (7%) | 11 (52%) | 41 (25%) | All pairs *p*<0.01 |
| Sputum | 446 (66%) | 199 (66%) | 108 (57%) | 20 (95%) | 119 (72%) | All pairs *p*<0.05 |
| Facial blushing | 273 (40%) | 135 (45%) | 33 (17%) | 12 (57%) | 93 (56%) | All pairs *p*<0.01 |
| Post-tussive vomiting | 202 (30%) | 101 (33%) | 32 (17%) | 8 (38%) | 61 (37%) | All pairs *p*<0.05 |
| Runny nose | 251 (37%) | 116 (38%) | 63 (33%) | 10 (48%) | 62 (38%) |  |
| Fever history | 72 (11%) | 38 (13%) | 18 (9%) | 3 (14%) | 13 (8%) |  |
| Apnea | 4 (1%) | 2 (1%) | 1 (1%) | 0 (0%) | 1 (1%) |  |
| Hospitalization | 138 (20%) | 81 (27%) | 12 (6%) | 8 (38%) | 37 (22%) | All pairs *p*<0.01 |
| Hospitalization days | 6 (5-10) | 6 (5-10) | 4 (1-7) | 6 (5-10) | 7 (5-10) | *ptxP1*: *p*<0.01<br>*ptxP3*-other: *p*<0.05 |
| **Chest computed tomography (CT)** |  |  |  |  |  |  |
| Normal | 159 (23%) | 64 (21%) | 59 (31%) | 2 (10%) | 34 (21%) | All pairs *p*<0.05 |
| Moderate (bronchitis) | 294 (43%) | 119 (39%) | 90 (47%) | 7 (33%) | 78 (47%) |  |
| Severe (bronchopneumonia or pneumonia) | 225 (33%) | 119 (39%) | 41 (22%) | 12 (57%) | 53 (32%) | All pairs *p*<0.05 |
| **Laboratory testing** |  |  |  |  |  |  |
| Abnormal CRP | 78 (12%) | 25 (8%) | 37 (20%) | 1 (5%) | 15 (9%) | *ptxP1*: *p*<0.01<br>*ptxP3*-other: *p*<0.01 |
| Abnormal WBC | 380 (58%) | 177 (61%) | 86 (47%) | 15 (71%) | 102 (64%) | All pairs *p*<0.05 |

Data are n (%) or median (IQR). CRP: C-reactive protein, WBC: white blood cell

Two phases of pertussis epidemic were identified in Shanghai, with the emergence of COVID-19 as transition point (figure 1). From pre- (n=447) to post-COVID-19 (n=845), the proportion of MRBP infections in children increased from 60% to 98%. Moreover, patients shifted from being dominant by infants (90%, 397/442) to widespread infection among older children (infant: 16%, 132/844), with a notable increase in the proportion of vaccinated population, surging from 31% (107/340) to 88% (664/756).

**Figure 1.**
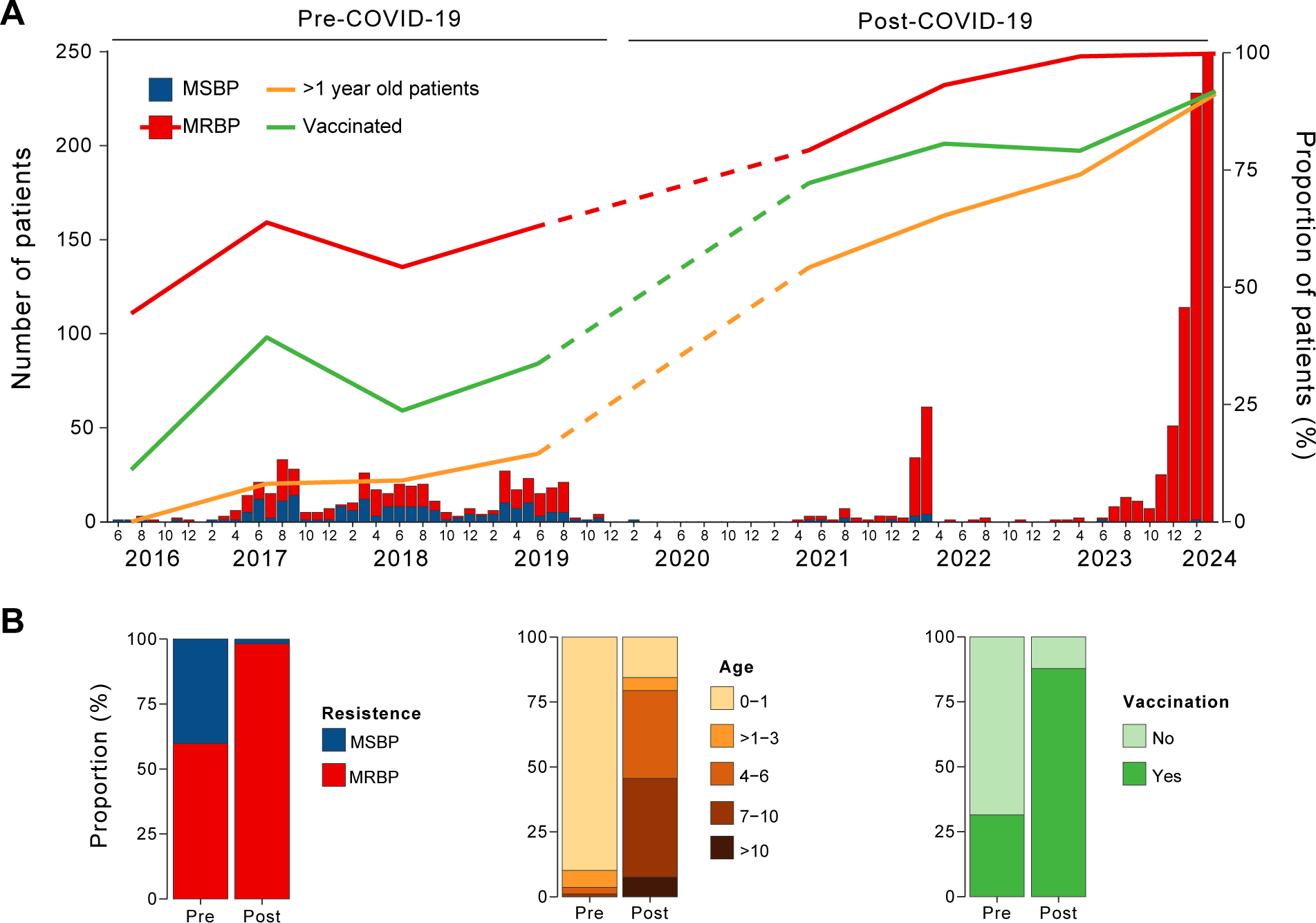
Pertussis epidemic dynamics in Shanghai, China between 2016 and 2024. (A) Monthly number of culture-positive children patients and annually proportions of MRBP/*ptxP3*/*ptxP3*-MRBP infection, older children (aged >1y) and vaccinated populations. Data from 2020 was not analysed due to insufficient sample size (n=1). (B) Transition of pertussis epidemiology from pre- to post-COVID-19 stage.

To characterize the circulating *B. pertussis* lineages, we constructed a phylogenic tree of 712 Shanghai strains based on core-genome SNPs (figure 2A). Two lineages were identified, which were designated as *ptxP1-* and *ptxP3*-lineage based on *ptxP* alleles. Nearly all (98%, 313/321) *ptxP1-*lineage strains and 50% (194/391) *ptxP3-*lineage strains were MRBP.

**Figure 2.**
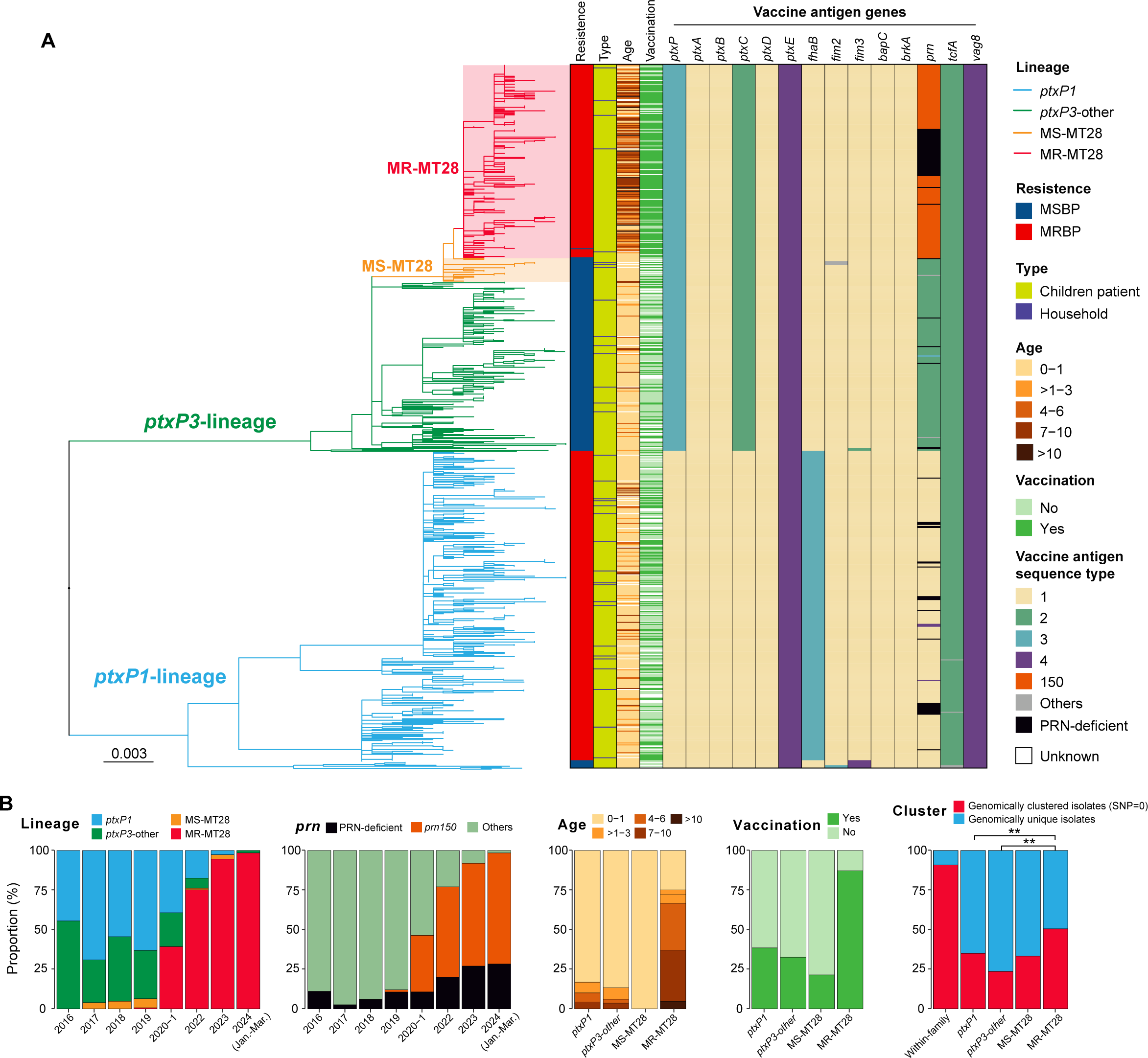
Phylogenomic and epidemiological characteristics of MR-MT28. (A) Phylogenetic tree, source and antigen gene alleles of Shanghai strains. (B) Temporal dynamics and age and vaccination composition of patients of different lineages/clones. (C) Proportion of genomically clustered strains.

Within *ptxP3-*lineage, all MRBP strains and one MSBP strain clustered together and belonged to a specific clone with MLVA type 28 (MT28), which was designed as MR-MT28. A novel antigen allele *prn150*, with one synonymous mutation (C531T) compared with *prn2*, was identified in MR-MT28. The *prn* of 26% (50/195) MR-MT28 strains were predicted to be PRN-deficient. The genotypes of MR-MT28 were *ptxP3/prn150* or PRN-deficient */fhaB1/ptxC2/ptxA1/firm2-1/fim3-1*. Except MR-MT28, all other MT28 strains were MSBP and designated as MS-MT28, which is closely related to MR-MT28 on the phylogenic tree (figure 2A). Non-MT28 strains of *ptxP3-*lineage was designated as *ptxP3-*other.

The emergence and expansion of MR-MT28 temporally strongly correlated with the transition of pertussis epidemiology and upsurge of cases (table 1 and figure 2B). Post-COVID-19, MR-MT28 proportion increased sharply and dominated infections since 2022. The proportion of *prn150* or PRN-deficient strains kept increasing alongside the expansion of MR-MT28. Notably, MR-MT28 uniquely caused widespread infection among older children (infant: 25%, 47/189) and accounted for a substantial proportion (87%, 150/172) of infections among vaccinated population. In contrast, other types predominantly infected infants (≥83%), with a much lower proportion of infections among vaccinated population (≤39%).

Increased transmission may contribute to the expansion of MR-MT28. To investigate this possibility, genomic clustering analysis based on SNP-distance was performed to infer putative transmissions. Strains from patient-household pairs were most likely from recent transmission. There was no SNP between strains from 30 (90%) of 33 patient-household pairs, which were consistent with recent transmission, while the remaining three pairs had 32 or more SNPs. We therefore selected zero SNP difference as the cutoff to infer genomic cluster representing putative transmission. 51% (96/190) MR-MT28 strains were assigned into genomic clusters, which was higher than other types and significantly (*p*<0·01) higher than *ptxP1* and *ptxP3*-other (figure 2C), indicating a potentially increased transmissibility of MR-MT28.

We randomly selected eight strains (MR-MT28: n=3, MS-MT28: n=3, *ptxP3*-other: n=2) for transcriptomic sequencing to investigate the possible molecular mechanism of MR-MT28 expansion. We found substantial differences of transcriptomic profiles between MT-28 (MR-MT28 and MS-MT28) and *ptxP3*-other strains (figure 3A), with 879 differentially expressed genes (DEGs) identified (appendix p 163). Enrichment analysis of DEGs indicated that virulence-related pathways including bacterial secretion system (ko03070) and pertussis (ko05133) were significantly (*p*<0·01) up-regulated (fold change >2) and bacterial motility proteins pathway was significantly downregulated (fold change <-2) in MT-28 strains. More specifically, most antigen genes were significantly upregulated, including Ptx encoding genes (*ptxA* to *ptxE*). There was limited difference of transcriptomic profiles between MR-MT28 and MS-MT28, with only eight DEGs not associated with known virulence or antigen genes identified (figure 3B).

**Figure 3.**
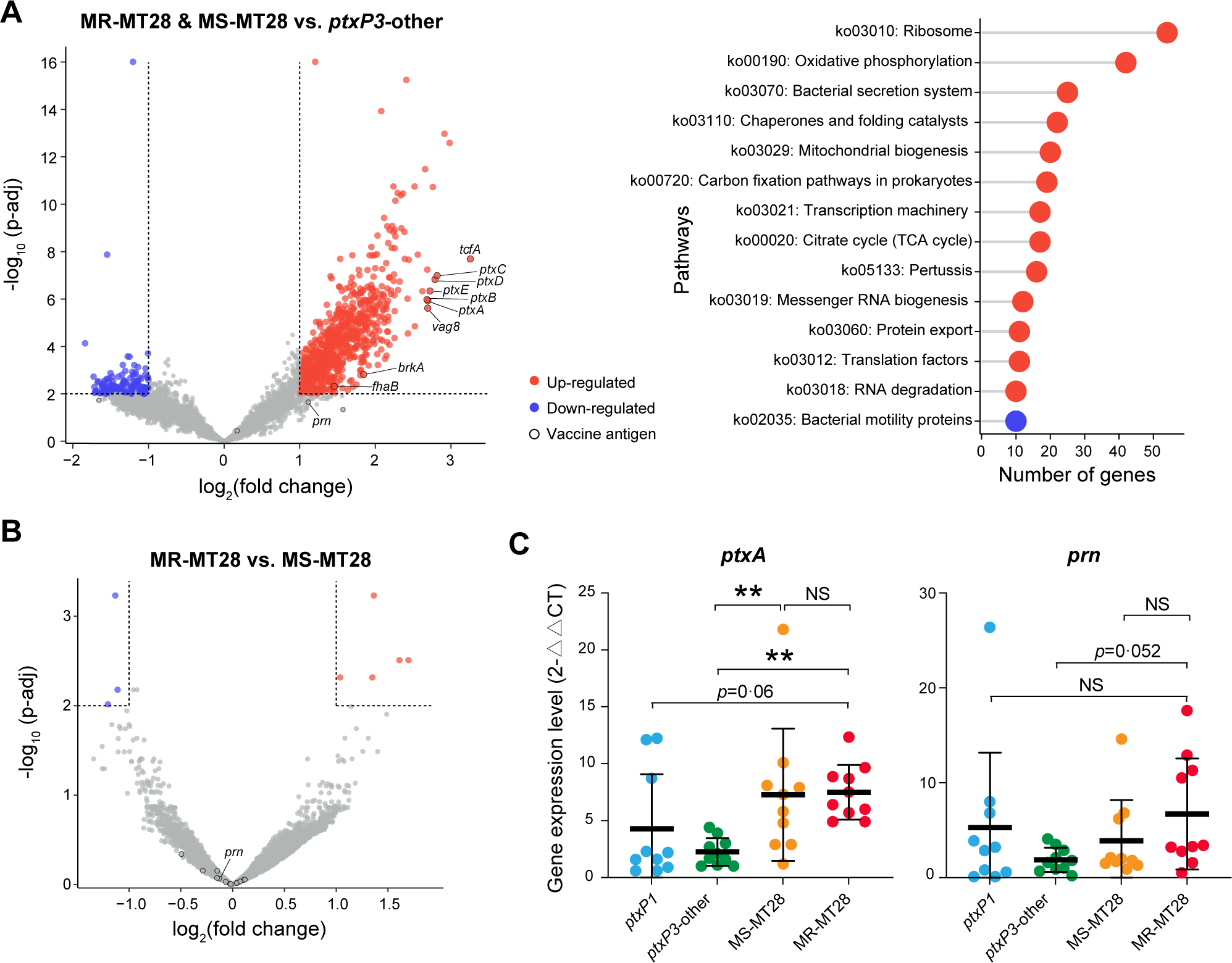
Transcriptomic (RNA-seq) and qPCR analyses of gene expression. (A,B) Volcano plot showing differentially expressed genes (left) and pathways (right). (C) Expression levels of seven antigen genes detected by qPCR. **: *p*<0·01, NS: not significant.

To validate gene expression results, we further quantified the expression of *ptxA* and *prn* of 40 strains (ten randomly selected strains per type) by quantitative PCR (qPCR, figure 3C). Consistent with transcriptomic analysis, the expression levels of *ptxA* in MT28 strains were significantly (*p*<0·01) higher than *ptxP3*-other strains and higher than *ptxP1*-lineage strains (*p*=0·06). There is no significant difference in the expression of *ptxA* and *prn* between MR-MT28 and MS-MT28 under the growth conditions tested, despite MR-MT28 carrying a novel *prn* allele.

In line with increased expression of Ptx encoding genes, the incidence of abnormal C-reactive protein (CRP) associated with MR-MT28 (20%) was significantly higher than *ptxP1*-lineage (8%) and *ptxP3*-other (9%), and higher than MS-MT28 (5%, *p*=0·089). However, overall, MR-MT28 infection caused milder clinical symptoms (table 1), including spasmodic cough, wheezing, sputum production, facial blushing and post-tussive vomiting, which were significantly (*p*<0·01 or 0·05) milder for MR-MT28 than other types. Moreover, MR-MT28 infections exhibited a significantly (*p*<0·05) higher proportion of normal chest computed tomography results and a significantly (*p*<0·01) lower hospitalization rate. Clinical symptoms among patients with same age catogory or vaccination status showed showed similar trends (appendix p1).

To investigate the origin and spread of MR-MT28 in China and globe, we newly sequenced 11 Beijing and Guangdong strains between 2019 and 2023, and compared with 6439 global strains. Phylogenetic analysis showed that MT28 clone existed in multiple regions worldwide, but so far, MR-MT28 was exclusively identified in China (figure 4A, B).

**Figure 4.**
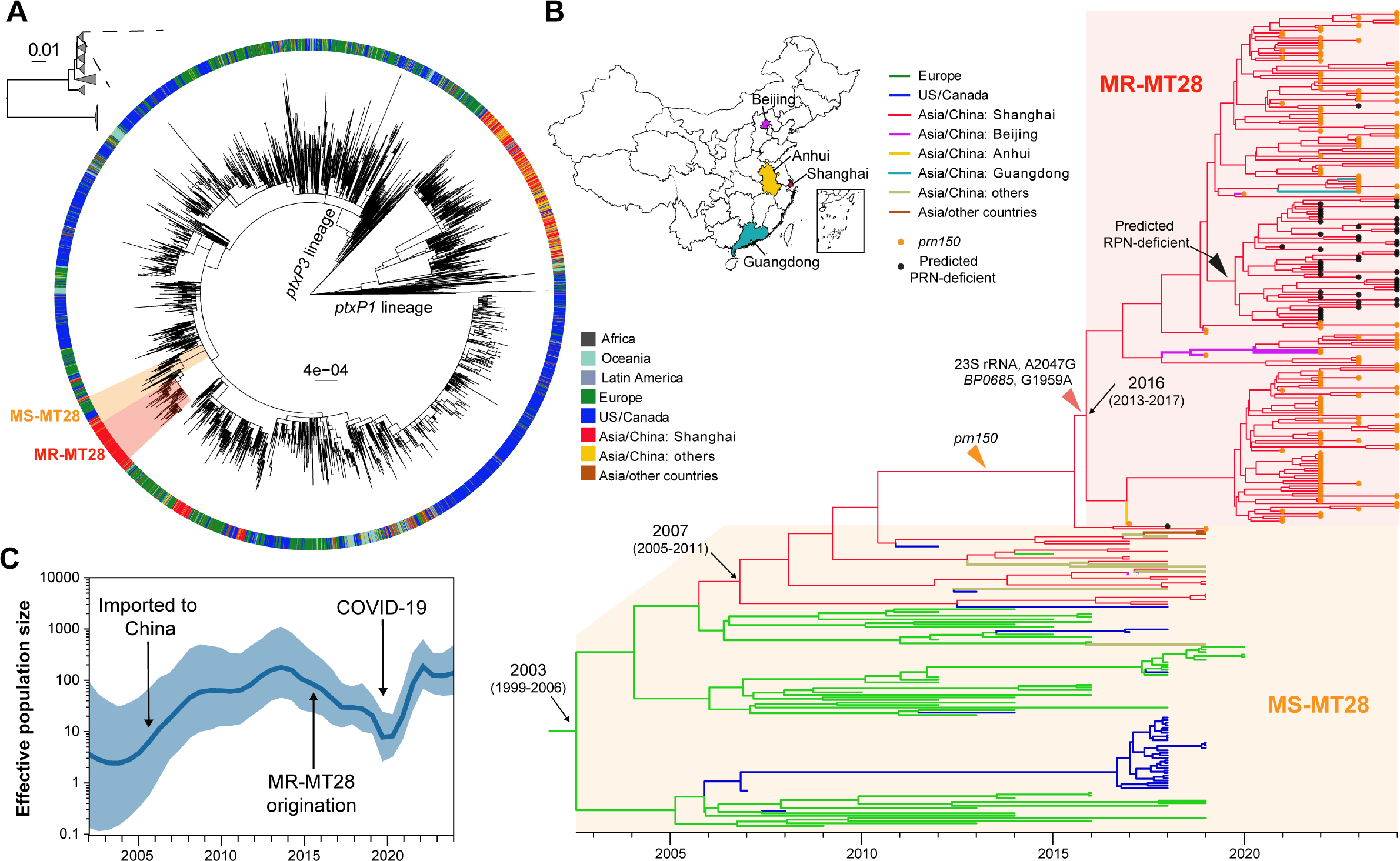
MR-MT28 in China and global context. (A) Phylogenic tree of Shanghai and global strains. The colors of outer ring indicated geographical regions. MR-MT28 and MS-MT28 were highlighted with red and orange backgrounds. (B) Maximum clade credibility tree of MT28 clone and geographical distribution of MR-MT28 in China. Branch colors indicated geographical regions. Key time points and genetic events were indicated by arrows and triangles. (C) Effective population size dynamics of MT28 clone inferred by Bayesian Skygrid analysis.

The MT28 clone was not a recently emerged one and likely originated from Europe around 2003 (95% highest probability density [HPD]: 1999-2006, figure 3B). It has spread in Europe and United States and was inferred to introduce to China around 2007 (95% HPD: 2005-2011). There are only two SNPs between MR-MT28 and MS-MT28 strains: 23S rRNA A2047G mutation and *BP0685* (dehydrogenase/oxidase) G15951A synonymous mutation. The *prn150* was not only identified in MR-MT28, but also found in a MS-MT28 strain closely related to MR-MT28 (figure 2A, 4B). After the acquisition of *prn150*, *BP0685* G15951A mutation and 23S rRNA A2047G mutation, MR-MT28 most likely originated in China in 2016 (95% HPD: 2013-2017), one year prior to the first report of MR-MT28 strain in Anhui, China in 2017.^21^ Notably, a sub-clone of MR-MT28 evolved into predicted PRN-deficient around 2019 (95% HPD: 2017-2019). Now MR-MT28 has been identified in four regions of China, including Anhui, Shanghai, Beijing and Guangdong (figure 4B). Beyond Shanghai, continuously identification of MR-MT28 occurred in Beijing during 2019-2022, indicating that domestic spread and colonization has occurred.

The effective population size of MT28 clone has steadily increase since its introduction into China (figure 4C). Intriguingly, following the origination of MR-MT28 in 2016, the overall population size did not increase but instead declined until the advent of COVID-19. Post-COVID-19, MR-MT28 population size increased rapidly, indicating that COVID-19 might be a critical factor for its rapid expansion.

## Discussion

The continuous surveillance and sampling in Shanghai provide a valuable opportunity to comprehensively investigate the link between pertussis upsurge, epidemiological transition, and *B. pertussis* evolution. Though the integration of large-scale WGS with phenotypic, transcriptomic, and clinical data, we identified and characterized a novel *ptxP3*-linage, macrolide-resistant clone, MR-MT28, which accounted for a substantial proportion of infections in older children and vaccinated individuals. The emergence and rapid expansion of MR-MT28 temporally strongly associated with pertussis upsurge, age shift, vaccine escape and sharp increase in MRBP prevalence.

While previous reports have noted pertussis age shift to older children and adolescents and vaccine escape,^4,5^ the situation in Shanghai exhibits distinct features. First, unlike gradual shifts observed in most countries, the shift in Shanghai occurred very quickly. Within two years, the proportion of older children (>1y) and vaccinated individuals surged from 10% and 31% to 84% and 88%, respectively, with the emergence of COVID-19 as the transition point. Second, pertussis shift in other countries was generally associated with the expansion of more virulent *ptxP3* strains, resulting in more severe clinical symptoms and higher hospitalization rates^16^. However, although MR-MT28 exhibited increased *ptxA* expression levels and higher incidence of abnormal CRP, it has been associated with generally milder clinical symptoms and a low hospitalization rate. Third, whereas the circulating *ptxP3*-lineage in most countries was polyphyletic with multiple subtypes and no dominant clone reported, a single clone, MR-MT28, dominated infections in Shanghai post-COVID-19 and has spread to multiple regions of China.

The causes of pertussis upsurge, age shift and vaccine escape have been attributed to multiple factors, including wanning immunity, ‘immunity debt’,^9^ improved diagnostics and pathogen evolution, with the first two being considered as the primary factors. ^2–5^ While wanning immunity or ‘immunity debt’ could explain the increased incidence among older children and upsurge of cases, it should not affect *B. pertussis* population composition and dynamics, and cannot explain the observation of MR-MT28 replacing other lineages and dominating infections. Improved diagnostics similarly do not affect *B. pertussis* population dynamics, and our results are based cultured strains, which are independent of diagnostic methods. Moreover, independent observations in Beijing^24^ provide further evidence that this scenario is not a bias caused by diagnostics.

We proposed that pathogen evolution is more likely the primary factor driving the scenario in Shanghai and China. MR-MT28 is uniquely capable of causing a substantial proportion of infections among older children and vaccinated individuals, suggesting enhanced vaccine escape. Moreover, the relatively milder clinical symptoms may prolong the interval before seeking medical care, thereby amplifying transmission opportunities. Phylogenomic clustering analysis provided evidence for the increased transmissibility of MR-MT28. Consequently, MR-MT28 may have competitive advantages due to antimicrobial resistance, enhanced vaccine escape, increased opportunities for transmission and transmissibility. In addition, the effective population size of MR-MT28 increased rapidly post-COVID-19, which could be related to the increased family contacts, particularly with unvaccinated infants, the major source for pertussis transmission,^29^ due to long-term lockdown and home life. These factors combined may have promoted the rapid expansion and dominance of MR-MT28.

While the molecular mechanism underlying *B. pertussis* vaccine escape remains unclear, three antigen genes have been implicated in this phenomenon: autotransporters genes *brkA* and *vag8*, and surface protein filamentous hemagglutinin gene *fhaB*.^30–33^ Intriguingly, all three genes were up-regulated in MR-MT28 compared with *ptxP3*-other strains. This, together with upreguation of Ptx encoding genes and other unidentified factors, may contribute to its enhanced vaccine escape. Despite the capacity to infect vaccinated individuals, clinical symptoms of MR-MT28 infection appear to be mild. This suggests that while the vaccine may not entirely prevent infection, it can attenuate clinical symptoms, a phenomenon also observed in other pathogens such as SARS-CoV-2.^34^

By integrating public data and 11 newly sequenced genomes from Beijing and Guangdong strains collected after 2019, our results showed the domestic spread and colonization of MR-MT28. Newly sequenced strains were randomly selected, and no information including subtyping and antimicrobial resistance was known in advance. Notably, a surprisingly high proportion of these strains (Beijing: 5/8, Guangdong: 2/3) were MR-MT28, suggesting that MR-MT28 may have cryptically dominated infections in these regions, like in Shanghai, but had not been identified due to limited surveillance and analysis or associated with their milder clinical symptoms. In addition to the expansion in China, MR-MT28 potentially has a high global spread risk because its consistent *ptxP3* allele and epidemiology across many counties. Given its resistance to first-line drugs and potentially competitive advantages, MR-MT28 warrants comprehensively global surveillance and research efforts.

Extremely high prevalence of MRBP indicates that macrolides are no longer suitable as first-line drugs for pertussis treatment in Shanghai and even China. Sulfamethoxazole/trimethoprim could be a good replacement because all MRBP strains were susceptible to it. While traditional drug susceptibility testing replies on strain culture and is time-consuming, PCR testing targeting specific mutations such as 23S rRNA A2047G mutation carried by all MRBP strains, offers a supplement or alternative. For MR-MT28, *BP0685* G15951A mutation or *prn150* are candidate targets, though the latter may present some problems (one non-MR-MT28 strains carried *prn150*, 26% MR-MT28 were predicted PRN-deficient).

Our studies have several limitations. First, most samples used for strain culture were obtained after patients had received antimicrobial treatment, which may introduce bias to the incidence of MRBP. However, this issue is common and remained consistent throughout the sampling period. Second, due to the unavailability of Ptx antibodies in China, we could only investigate the expression levels. Nonetheless, high expression does not necessarily correspond to increased Ptx production and virulence. Finally, while we provided evidence of domestic spread and colonization of MR-MT28, due to the small sample size except Shanghai, the incidence of MR-MT28 in China requires further evaluation based on more samples, especially samples collected post-COVID-19.

In conclusion, a novel *B. pertussis ptxP3*-linage clone, MR-MT28, has emerged and dominated infections in Shanghai post-COVID-19. MR-MT28 may have competitive advantages due to antimicrobial resistance, enhanced vaccine escape, increased opportunities for transmission and transmissibility, which drove the pertussis upsurge, age shift, vaccine escape and sharp increase in MRBP prevalence. We reconstructed the evolutionary history of MR-MT28 and demonstrated its domestic spread and colonization. This novel clone potentially has a higher global spread risk because its consistent *ptxP3* allele and epidemiology across many counties, which warrants further comprehensive global surveillance and research efforts. Macrolides may no longer be suitable as first-line drugs for pertussis treatment in China due to the extremely high prevalence of MRBP post-COVID-19.

## Data Availability

All data produced in the present study are available upon reasonable request to the authors

## Contributors

CW, PF, CY, and LG contributed to the conception of this project. PF and GY were responsible for collection of clinical and laboratory data. WC and PF performed clinical and laboratory data analysis. CY and LX performed bioinformatics analysis of whole genome sequence data. JQ, JZ and YL participated the genome preparision and qPCR. JQ, YK, SQ, SW, JZ and YL participated in the experiments. CY and PF prepared the manuscript. All authors contributed to the interpretation of results and critical review of the manuscript.

## Acknowledgments

This work was supported by grants from the National Key Research and Development Program of China (2021YFC2701800 and 2022YFC2304700), National Natural Science Foundation of China (82202567 and 32270003), Youth Innovation Promotion Association, Chinese Academy of Sciences (2022278), and Shanghai Rising-Star Program (23QA1410500), and Shanghai municipal three-year action plan for strengthening the construction of the public health system (2023-2025) GWVI-2.1.2.

